# Insights into spray impingement on mask surface: effect of mask properties on penetration and aerosolization of cough droplets

**DOI:** 10.1101/2021.11.27.21266925

**Authors:** Gautham Vadlamudi, S K Thirumalaikumaran, Dipshikha Chakravortty, Abhishek Saha, Saptarshi Basu

## Abstract

The emergence of the COVID-19 pandemic has demonstrated the importance of face masks, making them a part of people’s routine during the pandemic which is still continuing. The face masks act as source control, reducing the transmission of infectious respiratory droplets by acting as a physical barrier blocking the droplets during speaking, breathing, coughing, sneezing, etc. The novelty of current study is to generate a spray with the droplet size distribution and velocity scale similar that of an actual cough or a mild sneeze to fundamentally investigate the effects of mask properties on model-cough impingement. The spray replicates the presence of both large-sized and small-sized droplets similar to an actual cough, which makes the observations relevant to real-life situations. The spray is impinged on different mask samples with varying properties like porosity, pore size, fabric thickness, and their combinations in multilayer configuration. The effect of mask properties on the droplet penetration volume is studied as it leads to the release of higher pathogen loading into the surroundings. A two-step penetration criteria based on viscous dissipation and capillary effects have been applied along with a third criteria based on the porosity of the mask sample that is specifically applicable for the spray impingement. The droplets present in the impinging cough can penetrate through the mask, atomizing into the aerosolization range and thus increasing the infection potential. Hence the effect of mask properties on the droplet size distribution as well as the velocity distribution of the penetrated droplets has been investigated using *in-vitro* experimental manikin model, which will be essential for estimating the range of infection spread. The filtration of virus-emulating nanoparticles as well as the fate of the penetrated respiratory droplets, with a susceptible person in the proximity, has also been investigated.

## Introduction

The transportation of pathogen-loaded respiratory droplets from an infected person to a susceptible person can potentially result in triggering a global pandemic like the COVID-19^1-3^. The respiratory droplets will be ejected from an infected person during breathing, talking, singing, spitting, coughing, or sneezing, which can contribute to the spread of the virus or pathogen loaded droplets into the surroundings. These droplets when ingested into oral or nasal passages of a healthy person can lead to infection^4^. In case of larger size droplets (>100 μm), the ejected droplets can settle on the surfaces as fomites^5,6^ following the ballistic trajectory, exhibiting a shorter air borne lifetime represented using Wells curve^7^. The intermediate sized droplets may get carried over significant distances, and the smaller droplets (5-10 μm) remain suspended in air for longer durations. These smaller droplets may evaporate completely forming droplet nuclei or fomites which are transported from place to place by air flow or breeze^8,9^. The extent of spread and the infection probability due to the ejected droplets depends on the droplet size and ambient conditions. Chaudhuri et al. estimated that the droplets in the range of 10-50 μm have high infection potential and blocking all droplets above 5 μm using a face mask and social distancing is advised for restricting the spread of highly infectious diseases such as COVID 19^10,11^.

The face masks primarily contribute as a source control mechanism, and act as a physical barrier to obstruct the release of infected respiratory droplets into the surroundings^12^. Maclntyre et al. conducted clinical trial of face mask usage tracking the spread of different respiratory viruses and concluded that there is a relative reduction of about 60-80% in transmission with adherent mask usage^13^. Another important aspect of the study is that even with different transmission mechanisms like large droplet, aerosol or fomite, all the different major viruses are transmitted through respiratory route, which can be reduced with mask usage. However, the relative effectiveness depends on the type of the mask and properties of the mask used which reduces the distance travelled by the aerosolized droplets during human cough^14^. Konda et al. have carried out the investigation to find the filtration efficiencies of several common fabrics including cotton, silk, chiffon, flannel, various synthetics, and their combinations in multi-layered configurations using polydisperse NaCl aerosols^15^.

Droplet impact studies have been conducted by multiple researchers, using porous networks like metallic wire meshes, fibers, textiles and showed that the penetration characteristics depend on surface wettability, impact velocity, mesh size, and fluid properties^16-19^. However, Sahu et al. showed that beyond a critical impact velocity, penetration will occur through fiber pores irrespective of wettability^20^. Kooij et al. studied the ligament breakup and droplet fragmentation after impacting with a mesh and showed that the breakup is controlled by the instability arising due to initial perturbations in the droplet^16^. Krishan et al. have conducted experiments of droplet impingement on cotton masks, surgical masks and showed that the fabric properties like pore size and porosity affect the droplet penetration criteria^21^. Sharma et al. focused on the effect of number of layers present in a surgical mask and showed that volume of daughter droplets was significantly reduced after penetration, due to the additional layer^22^. Bagchi et al. have investigated the droplet impingement on wet facemasks to study the penetration and secondary atomization phenomena^23^.

Arumuru et al. have determined the average distance travelled by the sneeze droplets with different combinations of face masks and face shields using jet visualization^24^. Verma et al. have investigated the efficacy of different commercially available face masks by determining the distance travelled by the replicated respiratory jets with different type of masks^25^. Many investigations have been conducted by researchers to test the efficacy of masks, all of which primarily focused on smaller sized droplets (0-100 μm)^14,26-29^. Flow field generated by coughing with and without surgical mask has been investigated by Kahler et al. to study the flow blockage caused by the masks using PIV measurements^29^. Optical Schlieren experiments conducted by Tang et al. showed that the forward jet of droplets during a cough event are blocked by the mask, however leakage around the top, bottom and sides is observed^28^. Manikin experiments are done by Pan et al. to evaluate the inward and outward effectiveness of cloth masks, surgical masks, face shield using aerosol generators. The filtration efficiency has been measured using aerodynamic particle sizer spectrometer connected to the inhaling manikin. They recommended three-layered mask consisting of outer layers of a flexible, tightly woven fabric and inner layer consisting of a material designed to filter out particles^30^. Rothamer et al. have also performed manikin studies to evaluate the impact of ventilation on aerosol dynamics in a classroom setting and the aerosol was found to be present in the room uniformly for a distance of 2m. They have studied the filtration efficiency of different types of masks. It showed that high filtration efficient masks are necessary to reduce the infection probability, as the effect of ventilation alone is not enough to control the spread^31^. The effectiveness of blocking of the virus-like nanoparticles by common fabrics has been investigated by Lustig et al.^32^. The mask efficiency, leakage and filtration for multiple cough cycles during mild cough has been investigated by Dbouk et al.^33^.

It is reported that the saliva droplets ejected from a human cough can travel 2 m of distance with zero wind speed and the range may increase to 6 m with wind speed of 4-15 kmph^34^. Studies have been conducted by Lindsley et al. using cough simulators which can generate air flow of 32 m/s having cough aerosol particles (0-100 μm)^35^. It is to be noted that the respiratory droplets during a cough are characterized by wide range of sizes, from submicrometers to few millimeters^9,28,36^ with an average velocity of 10 m/s^37,38^. Even though the number of large-sized droplets (> 250 μm) is small in a cough, a significantly large percentage of total volume (94.21%) is contributed by these larger droplets^9^. Duguid has shown a direct correlation between the amount of pathogen loading with the droplet volume. This suggests that these larger sized droplets are also relevant in disease transmission along with the aerosolized droplets (< 100 μm) having longer air borne lifetime. Furthermore, Yan et al. have shown that large droplets will also be produced during both sneezing and coughing, along with the aerosol droplets^39^.

Most of the afore-mentioned experimental investigations on mask involved the study of flow field and aerosolized jets at similar Reynolds number associated with the actual cough or sneeze. Majority of these studies focused on the distance travelled by the droplets, its correlation with different type of masks or effect of mask fit or ventilation etc. The individual droplet impact studies in the literature were primarily focusing on the larger droplets and their penetration criteria on different meshes or fabrics. However, as discussed before, both larger droplets and smaller droplets are produced during a coughing or a sneezing event and there is very sparse literature on model-cough droplet impingement using a spray. The spray having the droplet size range of 0-500 μm impinging on a mask surface with velocity of ∼10 m/s closely resembles the characteristics of an actual cough or a mild sneeze which is rarely studied in the literature. A fundamental study on this spray impingement gives insights into the droplet penetration criteria, possibility of secondary atomization, ligament formation, ligament breakup and size distribution of the daughter droplets. Furthermore, it gives insight into how the mask properties like porosity, pore size and number of layers affect these phenomena. In our previous studies, the single droplet impingement has been studied to get insight into the penetration phenomenon locally at each droplet^21,22^. In continuation, experimental investigation has been conducted in current study replicating a model-cough by impinging a spray having similar droplet size and velocity range associated with actual cough. The effect of different fabric properties of mask samples on penetration dynamics has been addressed. Based on the spray impingement experiments, the effect of penetration on different sized droplets present in the spray of a cough is also investigated.

Further, the filtration of virus-emulating nanoparticles investigated along with the fate of penetrated respiratory droplets in the presence of a susceptible person in proximity.

## Results and discussions

### Droplet penetration criteria

#### Two step penetration criteria

A penetration-criteria have to be developed for determining whether the penetration of the droplets occurs through the pores of the mask or not. Following in similar lines as in our previous studies and the literature^21,22^, the main physical mechanisms which oppose or resist the penetration of droplets through the mask are capillary forces due to surface tension and viscous dissipation at the pore-level.

Hence, the penetration criteria have to consider both opposing effects i.e., one with respect to viscous dissipation (Reynolds number, *Re*_*ϵ*_) and other with respect to surface tension effects (Weber Number, *We*_*ϵ*_). Since both these effects oppose the penetration of droplets, the momentum in the impinging droplets has to overpower both capillary and viscous dissipation effects for the penetration to be possible. Hence, two step criteria is considered based on capillary effects and viscous dissipation, both of which have to be followed for the penetration to occur. Sahu et al.^20^ reported that beyond a threshold impact velocity, irrespective of hydrophobicity and initial mass, the liquid penetration always occurs through a porous network of fibers. The liquid passes through the pores of the fabric in the form of ligaments, extruding outward from the pores. For a fabric pore size of *ϵ* and thickness *t*_*m*_, the liquid ligaments can pass through the pores when the kinetic energy exceeds the viscous dissipation in the pores^20,21^. A penetration-criteria has been formulated using this argument based on viscous dissipation as given below:

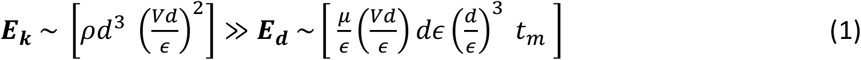

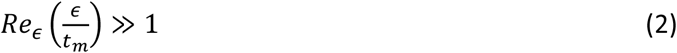

This suggests that penetration only occurs when the condition 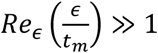 followed.

The second penetration criterion is based on the surface tension effects, which suggest that for droplet penetration can only occur when the dynamic pressure (∼ *ρV*^2^) exerted by the impinging droplet has to exceed the capillary pressure (∼ 4*σ*/ *ϵ*) inside the fabric pores^21^. Hence, according to the surface tension criteria, penetration only occurs when:

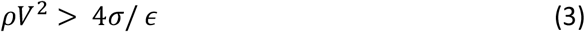

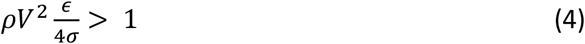

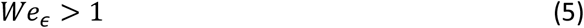

Where ρ is liquid density, σ is the surface tension between the water-air interface, V is impact velocity, ϵ is pore size and We is weber number.

These two criteria do not contain any droplet diameter dependent term and are only dependent on the mask fabric properties. However, it is to be noted that these criteria are derived under the assumption of *d* > ϵ and will not be valid for *d* < ϵ cases. Additionally, it is evident from the Eq.s [2, 5] that the two mask properties ***ϵ*** and *t*_*m*_ affect the two penetration criteria differently. Pore size (***ϵ***) affects both viscous dissipation as well as surface tension criteria, however thickness (*t*_*m*_) only effects the viscous dissipation criteria. Higher the value of *t*_*m*_, the parameter for viscous dissipation criteria will be lower, suggesting higher viscous dissipation. From this the relative importance of the two criteria in case of a mask sample with specific properties can be known by the ratio of *t*_*m*_ and ***ϵ***. Higher value of the ratio ***t***_***m***_/***ϵ*** for a given mask sample suggests that viscous dissipation criterion is more dominant and relevant in determining the possibility of droplet penetration (see Eq.s [2]). Considering the maximum value of velocity (∼14 m/s) associated with the impinging spray in current experiments, the value of parameters of the two criteria and the ratio ***t***_***m***_/***ϵ*** for all the different test samples has been tabulated in table 2. It is to be noted that the penetration phenomenon fundamentally occurs at the local pore-level. Hence, only the local properties of the mask like pore size (***ϵ***), thickness (***t***_***m***_) are considered for the penetration criteria but not porosity (***ϕ***) which is a global parameter and doesn’t affect the pore-level dynamics.

**Table 1:** The Characteristics of different mask samples and their combinations in multilayer configuration.

| Sl no. | Samples | Porosity ($\phi$) % | Pore size, $\epsilon$ ($\mu\text{m}$) | Thickness, $t_m$ ($\mu\text{m}$) |
| --- | --- | --- | --- | --- |
| 1 | Layer A | $17.22 \pm 2.72$ | $45.20 \pm 3.64$ | $430.11 \pm 12.568$ |
| 2 | Layer B | $0.14 \pm 0.01$ | $22.20 \pm 0.79$ | $595.26 \pm 13.96$ |
| 3 | Layer C | $4.84 \pm 0.78$ | $31.68 \pm 1.08$ | $512.7 \pm 12.195$ |
| 4 | 2 x Layer A | $5.23 \pm 0.77$ | $31.51 \pm 1.04$ | $873.42 \pm 11.759$ |
| 5 | Layer A + C | $1.95 \pm 0.31$ | $25.90 \pm 0.89$ | $950.39 \pm 11.152$ |
| 6 | 2 x Layer C | $0.33 \pm 0.02$ | $23.40 \pm 0.72$ | $1086.21 \pm 13.468$ |
| 7 | 3 x Layer A | $0.39 \pm 0.09$ | $24.03 \pm 0.84$ | $1285.3 \pm 15.661$ |
| 8 | 3 x Layer C | $0.04 \pm 0.01$ | $28.44 \pm 0.61$ | $1546.95 \pm 11.293$ |

**Table 2.** Table shows the two penetration criteria for different mask samples calculated at maximum value of impingement velocity (∼ 14 m/s). The values of porosity (***ϕ***), pore size (***ϵ***), thickness (***t***_***m***_) and ***t***_***m***_/***ϵ*** ratio are also tabulated corresponding to the respective samples. The two penetration criteria are based on **viscous dissipation:** 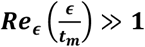, and **surface tension: *We***_***ϵ***_ > **1**. Green color fill indicates that penetration does not occur, and orange color fill indicates that penetration occurs, for those samples experimentally. Based on the corresponding criterion, red font indicates that penetration is possible and green font indicates that penetration is not possible, for that particular sample.

| | $\phi$ % | $\epsilon$ ($\mu\text{m}$) | $t_m$ ($\mu\text{m}$) | $Re_\epsilon \left( \frac{\epsilon}{t_m} \right)$ | $t_m/\epsilon$ | $We_\epsilon$ |
| --- | --- | --- | --- | --- | --- | --- |
| Layer A | 17.22 | 45.2 | 430 | 77.96 | 9.51 | 30.76 |
| Layer C | 4.84 | 31.68 | 513 | 32.10 | 16.19 | 21.56 |
| 2 x Layer A | 5.23 | 31.51 | 873 | 18.66 | 27.70 | 21.44 |
| Layer A + C | 1.95 | 25.9 | 950 | 11.58 | 36.67 | 17.63 |
| 2 x Layer C | 0.33 | 23.4 | 1086 | 8.27 | 46.41 | 15.93 |
| 3 x Layer A | 0.39 | 24.03 | 1285 | 7.37 | 53.47 | 16.35 |
| Layer B | 0.14 | 22.2 | 595 | 13.59 | 26.80 | 15.11 |
| 3 x Layer C | 0.04 | 28.44 | 1547 | 8.58 | 54.39 | 19.36 |

From the table 2, it can be observed that the ***t***_***m***_/***ϵ*** ratio is of order O (10) for all the cases, suggesting that the viscous dissipation criteria should be more relevant for these mask samples. This matches with the observation of Krishan et al. showing that the ***t***_***m***_/***ϵ*** ratio is of order O (10) in case of surgical mask which are used in the current study^21^. This is reflected in the surface tension criteria parameter values i.e., for all the samples, *We*_*ϵ*_ > 15 (far away from the criterion i.e., *We*_*ϵ*_ > 1), which suggests that the capillary pressure is overpowered by the dynamic pressure of impinging droplets in all the cases, making the surface tension effects negligible. This suggests that viscous dissipation is the only other resisting or opposing effect that can impede the penetration. This necessitates the reliance on viscous dissipation criteria as well, since the surface tension criteria alone is insufficient to estimate the penetration characteristics of the spray impingement. Using viscous dissipation criteria, the value of the parameter (*Re*_*ϵ*_ (*ϵ*/*t*_*m*_)) is found to be of order O (1) for the samples **2 x C, 3 x A**, and **3 x C**, and its value is ≫ **1** i.e., O (10) for all the other samples. This suggests that according to the viscous dissipation criteria, penetration should not occur in case of the three samples **2 x C, 3 x A**, and **3 x C**. However, from the experiments, the penetration is not observed only for the two samples **B** and **3 x C** samples. Additionally, in case of the sample **3 x A**, penetration observed is relatively very low. Hence, we can conclude that the viscous dissipation criterion is relatively in good agreement with the experiments in predicting the absence of penetration with minor discrepancy in case of the sample **3 x A**. However, experimentally the sample **B** does not exhibit penetration as well. This is not incorporated using the viscous dissipation criteria, a part of the two step criteria which is based on the local parameters. The local pore-level two step criteria as shown in this section, are able to predict the penetration possibility correctly for all the samples except one, which is reasonable. However, these two step criteria is only applicable fully for single droplet impingement and may not be entirely sufficient for spray impingement.

#### Additional penetration criteria for spray impingement

These two criteria based on viscous dissipation and surface tension, focus only on the penetration characteristics locally at the pore-level, which is the possible reason for this discrepancy. Since the current experiments involve spray impingement, the two step criteria do not give full picture about the penetration phenomena, as spray is not localized at an individual pore. In case of spray, the penetration may not always occur as the individual droplets might not always come across the gaps or pores to pass through, even though the local mask properties allow the penetration. Since, those criteria are focusing on local pore-level penetration, they may not be sufficient to predict extent of penetration for spray impingement with global outlook, without considering the global parameters. Hence, in addition to the pore-level two step criteria, a global property has to be considered in the scaling, especially in order to estimate the penetration for sprays.

Porosity (***ϕ***) is one such a property of mask, which is the proportion of total amount of space or gap available for droplets to penetrate through and it is not local pore-level property. In order to get a global penetration criterion, both global parameters corresponding to mask as well as the impinging spray are needed to be scaled together. Porosity (***ϕ***) can be considered as one of the parameters for global mask property and there is a necessity for the formulation of a global spray parameter which is analogous to porosity. Since, porosity is the cross-sectional areal ratio of available gaps to total area, a cross-sectional areal quantity has to be scaled for the impinging spray. Hence, a new spray parameter is formulated called cross-sectional spray areal density (ℂ) which is the proportion of total cross-sectional projected area of spray droplets (*A*_*spray c*/*s*_) with respect to the available area of the mask at any given instant, at the point of impact.

The side-view shadowgraphy images are used to estimate the parameter ℂ, due to the experimental limitations to accurately capture the cross-sectional view of the droplets in the impinging spray. As shown in the Fig. 1, the droplets present at the vicinity of the mask surface just before the impact are considered and their individual horizontal width is used for the calculation of projected area for each droplet assuming axisymmetry. The corresponding cross-sectional projected area summed over all the droplets that are located just above the mask, to calculate the total cross-sectional projected area of the spray (*A*_*spray c*/*s*_). This value is averaged over multiple frames and multiple experimental runs, in the window of consideration to get the final value of *A*_*spray c*/*s*_. The ratio of *A*_*spray c*/*s*_ and total mask cross-sectional area (*A*_*mask*_∼*h*^2^) gives the parameter ℂ (cross-sectional spray areal density) whose value is found to be (1.58 ± 0.3) %. The same averaged value of ℂ is used for all the cases for simplicity since the valve opening time and upstream pressure were maintained same for generating the spray in all the experimental runs for all the cases. Finally, for spray penetration criterion, the ratio of the two global parameters based on the mask (***ϕ***) and spray (ℂ) properties is considered, given as:

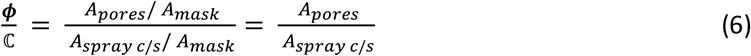

**Fig. 1.**
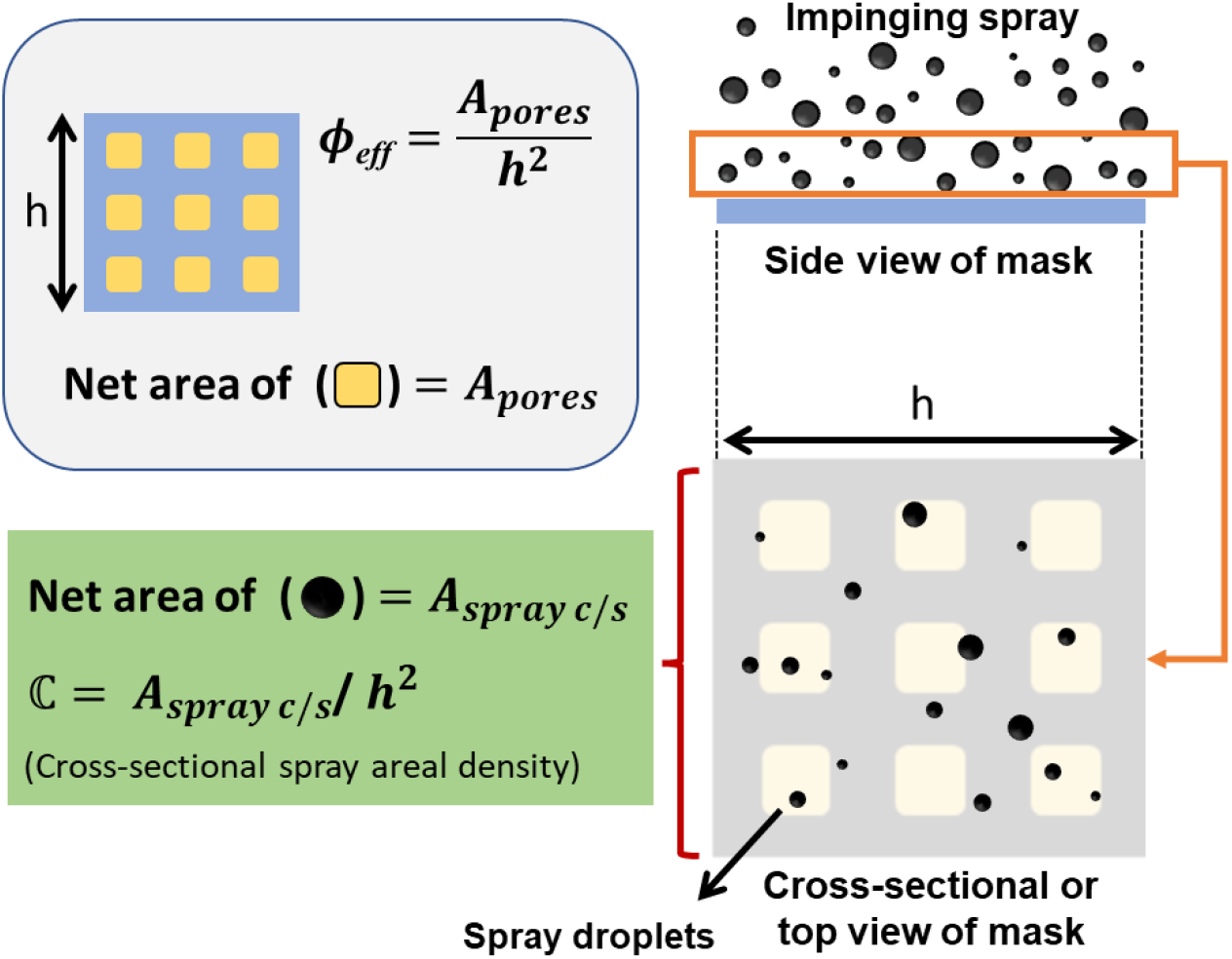
The methodology of calculation of the parameter ℂ (cross-sectional spray areal density) from experimental data. The spray droplets just before their impact on the mask (enclosed by orange box) are considered for the calculation of ℂ. Calculation of porosity is given in the top left.

The ratio ***ϕ***/ℂ represents the amount of pore area available per unit spray cross-sectional area. This gives a qualitative measure of the probability of penetration of the spray. Lower value of ***ϕ***/ℂ leads to lesser probability of spray penetration. Hence, for 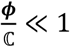, the probability of spray penetration is very less, leading to significantly low penetration volumes.

The discrepancy observed in case of two-step criteria is because the criteria do not account for the porosity of the sample (***ϕ***). The sample **B** shows no penetration experimentally but the two step criteria predicted penetration is possible in this sample. However, the significantly low value of porosity for this sample **B** (***ϕ*** ∼ **0. 14**) results in restricting the penetration even though penetration should be theoretically possible at the pore-level as the initial kinetic energy can surpass both viscous dissipation and capillary effects. This effect is incorporated using the ratio ***ϕ***/ℂ (shown in table 3) where the two samples **B** and **3 x C** which exhibit no penetration experimentally, has significantly low value of ratio ***ϕ***/ℂ in the order ≤ *O* (0.1), as shown in table 3. Hence, we can conclude that this third criterion suggesting penetration will not occur for 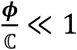 is in good agreement with the experiments.

**Table 3.**
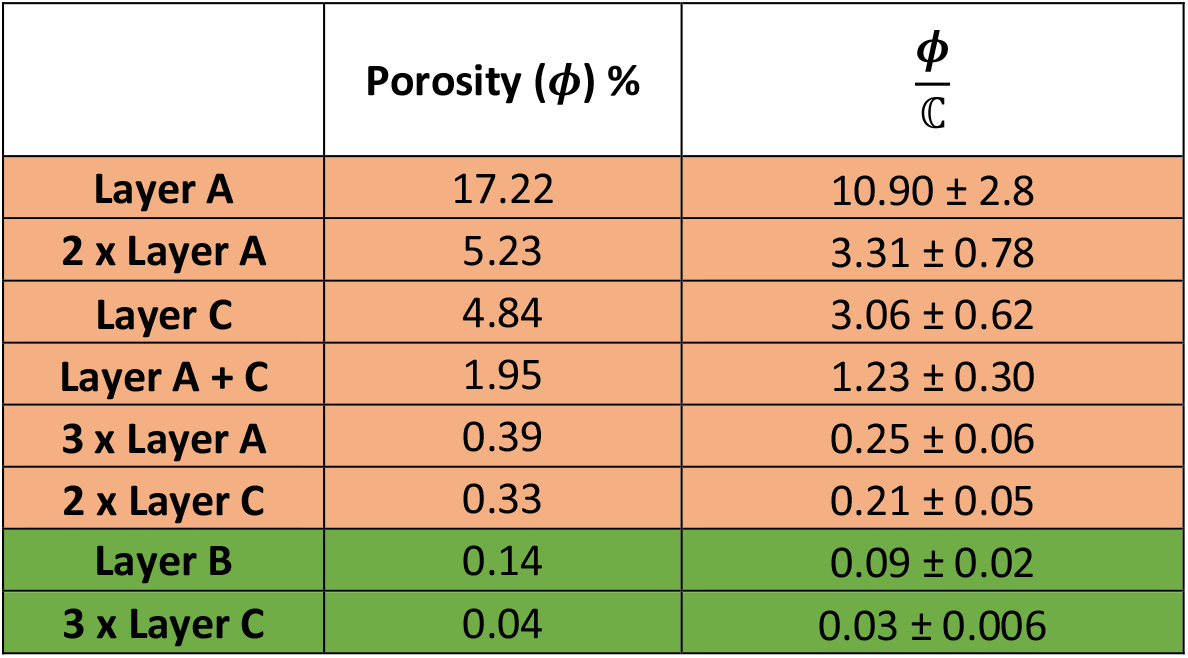
The table shows the values of the ratio ***ϕ***/ℂ in the decreasing order of the corresponding porosity (***ϕ***) for different samples giving a qualitative estimate for penetration probability for spray impingement. The green fill represents no penetration and orange fill indicates the occurrence of penetration for the respective sample.

Additionally, from the table 3 it can be observed that the values of the ratio 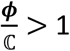 for the samples **A, C, A + C, 2 x A**, suggesting higher probability of spray penetration which agrees with the experiments. For the other samples in the range of 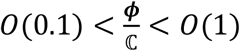 i.e., samples **A + C, 3 x A** and **2 x B**, the penetration is significantly lower due to lower value of the ratio ***ϕ***/ℂ. This newly defined ratio ***ϕ***/ℂ gives an estimate of the penetration possibility of the spray on a mask surface at global scale. Thus, the combination of the two step criteria and the ***ϕ***/ℂ ratio can give accurate penetration criteria for the spray impingement on a mask fabric, which agrees well with the experimental observations.

### Penetrated volume percentage

The droplet impingement on a porous surface like mask fabric involves three different phenomena^21^. The liquid passes through the fabric pores and extrudes out on the other side in the form of ligaments. These ligaments break up due to Rayleigh-Plateau instability forming daughter droplets. Some portion of initial droplet spreads along the fabric surface and bounce-back occurs. The remaining volume of the droplet is retained in the fabric through absorption and percolation. As the droplet impinges on the mask fabric, the initial kinetic energy of the impinging droplet is lost to viscous dissipation, overcoming surface tension, and spreading of liquid along the fabric. The remaining available energy is converted into the kinetic energy of the ligaments that extrude through the fabric pores. This available kinetic energy dictates the ligament length and velocity of the daughter droplets after the ligament break up. The size and velocity of the ligaments also effect the size of the daughter droplets.

The droplet penetration involved in the spray impingement is characterized by multiple droplet sizes and velocity scales and the dependency on droplet diameter on penetration needs to be investigated. The time series images of spray impingement and penetration for sample **A** has been shown in Fig. 2. The process of formation of ligaments due to extrusion of droplets through the pores can be observed in Fig. 2 (a) time series images for different impinging droplet diameters. The time series images clearly show the process of atomization of spray droplets into smaller size (aerosol range) at high impingement velocity. The zoomed-in version of larger droplet impingement in the spray (∼500 μm) has been shown in Fig. 2 (b), which clearly shows the formation of ligaments and ligament breakup phenomenon caused by Rayleigh-Plateau instability^21^. This ligament breakup leads to the formation of daughter droplets having lower diameter range, depending on the pore size and ligament velocity. Moreover, the zoomed-in images of the impingement of smaller droplets (∼100 μm) is shown in Fig. 2 (c), which clearly shows penetration of smaller droplet as well, given sufficient kinetic energy. These small droplets (diameter larger than pore size) undergo atomization into further smaller sized daughter droplets through ligament formation. Hence, it can be inferred that both larger and smaller droplets in the cough can potentially lead to the aerosolization and contribute to the volume-penetrated.

**Fig. 2.**
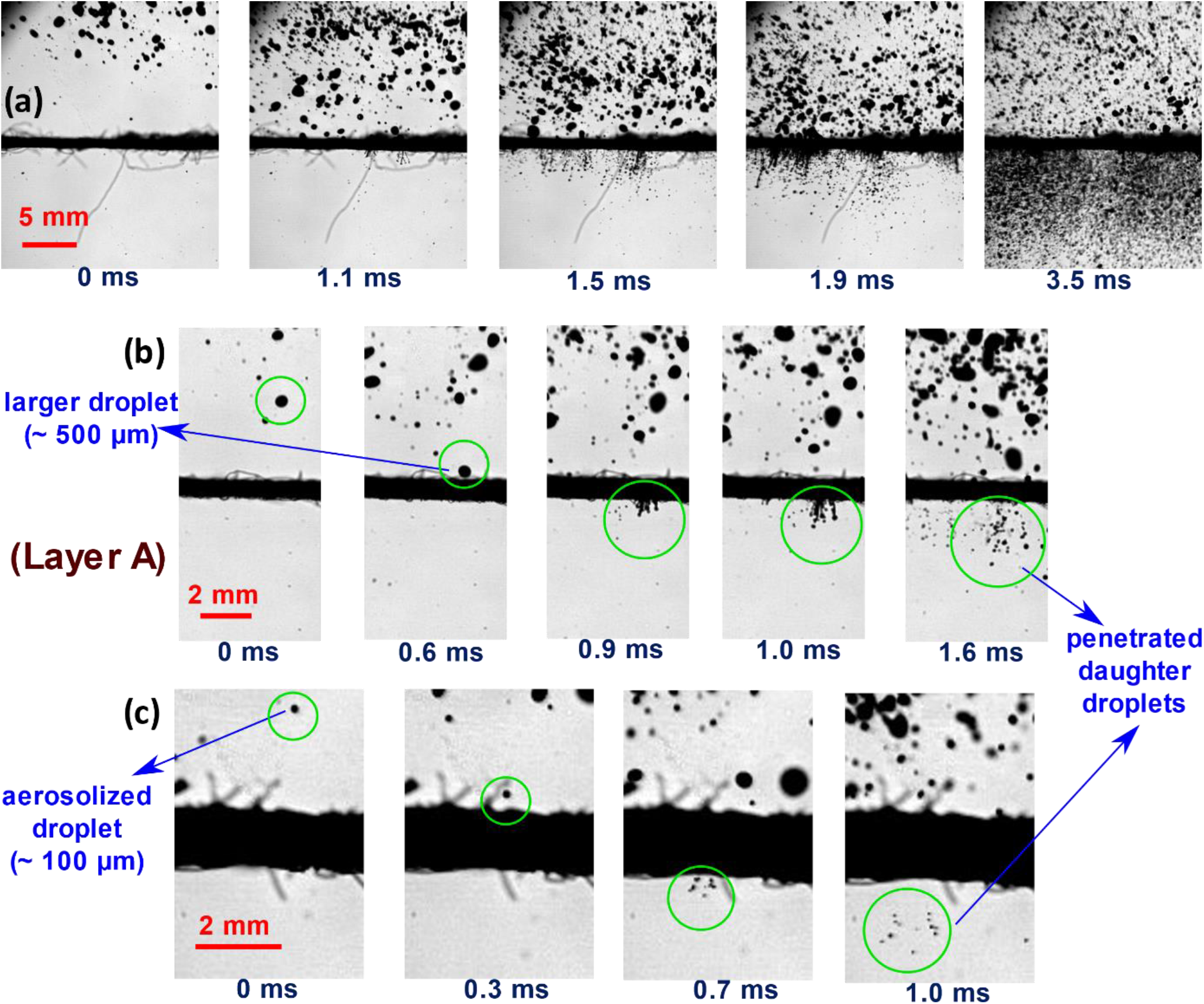
**(a)** The time series images of the spray impingement on a mask sample (Layer **A**). **(b)** Zoomed in time series images of impingement of larger sized droplet (∼500 μm) in the impinging spray (for sample **A**), **(c)** Zoomed in time series images of impingement of aerosolized smaller sized droplet (∼100 μm) in the impinging spray (for sample **A**). All the scale bars are represented in the images.

The percentage volume-penetrated during the spray impingement has been tabulated in the table 4. The percentage volume-penetrated is calculated from the shadowgraphy experiments and the calculated values are in good agreement with the microbalance measurements of the penetrated spray for different cases with the uncertainty less than ± 10 %. The pore size (***ϵ***) of all the samples is in similar range and their values are in same order of O (10), because of which ***ϵ*** does not have significant effect on the percentage volume-penetrated. However, the porosity (***ϕ***) effects the volume penetration percentage because the net gap available for the liquid to pass through directly affects the volume-penetrated.

**Table 4.** Table shows the different samples in decreasing order of the penetration volume percentage in spray impingement experiments. The values of porosity (***ϕ***), pore size (***ϵ***), thickness (***t***_***m***_) and ***t***_***m***_/***ϵ*** ratio are also tabulated corresponding to the respective samples. The penetration volume percentage generally follows a decreasing trend with the porosity. Blue color fill indicates the samples for which percentage volume-penetrated deviates from this trend. The red font indicates the lower penetration volume percentage in the deviating samples due to higher thickness. This effect of ***t***_***m***_ is attributed to higher ***t***_***m***_/***ϵ*** ratio indicating the dominance of viscous dissipation effects. Green fill indicates no penetration, and the orange fill indicates occurrence of penetration for the respective sample.

| | $\phi$ % | $\epsilon$ ($\mu\text{m}$) | $t_m$ ($\mu\text{m}$) | $t_m/\epsilon$ | % Volume-penetrated |
| --- | --- | --- | --- | --- | --- |
| Layer A | 17.22 | 45.2 | 430 | 9.51 | 16.67 |
| Layer C | 4.84 | 31.68 | 513 | 16.19 | 4.55 |
| 2 x Layer A | 5.23 | 31.51 | 873 | 27.70 | 0.85 |
| Layer A + C | 1.95 | 25.9 | 950 | 36.67 | 0.38 |
| 2 x Layer C | 0.33 | 23.4 | 1086 | 46.41 | 0.34 |
| 3 x Layer A | 0.39 | 24.03 | 1285 | 53.47 | 0.20 |
| Layer B | 0.14 | 22.2 | 595 | 26.80 | 0.00 |
| 3 x Layer C | 0.04 | 28.44 | 1547 | 54.39 | 0.00 |

Additionally, a general trend is observed where the percentage volume-penetrated decreases with decrease in porosity (***ϕ***), however there are some exceptions to this general trend. In case of the samples **C** and **2 x A**, the porosity increases slightly from samples **C** to **2 x A**, but the percentage volume-penetrated decreased drastically. This is because of the effect of significantly higher fabric thickness (***t***_***m***_) in sample **2 x A** compared to sample **C** and both the samples having the porosity (***ϕ***) and pore size (***ϵ***) in relatively similar range. The larger value of fiber thickness (***t***_***m***_) contributed to higher viscous dissipation leading to lower penetrated volume percentage in sample **2 x A** compared to **C**. In case of the samples **2 x C** and **3 x A**, the similar to the previous case, the porosity (***ϕ***) and pore size (***ϵ***) are in similar range, however fabric thickness (***t***_***m***_) is relatively higher in case of **3 x A**. Moreover, the effect of the increase in fabric thickness (***t***_***m***_) is more compared to the effect of the increment in porosity (***ϕ***) or pore size (***ϵ***), which caused the net decrease in percentage volume-penetrated from sample **2 x C** to sample **3 x A**. The dominant effect of the fabric thickness (***t***_***m***_) on penetration volume can be attributed to the higher value of ***t***_***m***_/***ϵ***, suggesting the dominance of viscous dissipation effects in these samples as mentioned in the previous section. Except for the samples **B** and **3 x C**, all the other samples exhibited penetration. Another interesting observation is that the penetration volume percentage has a weak positive correlation with the ***ϕ***/ℂ ratio, because of the porosity variation. It is to be noted that the ***ϕ***/ℂ ratio gives a decent estimate of penetration of the spray. For ***ϕ***/ℂ < 1, all the samples have significantly low penetration volume percentage (< 0.4 %) suggesting that ***ϕ***/ℂ ratio is decently effective in giving rough estimate of extent of penetration. Additionally, the ***ϕ***/ℂ ratio accurately predicts the possibility of penetration in the samples in which the local mask properties allow penetration but the global property (***ϕ***) may or may not always allow the penetration depending on the impinging spray density which is incorporated in ***ϕ***/ℂ ratio. The two-step criteria gives insight into the possibility of penetration, however in case of spray impingement the complete picture is only obtained when it is combined with ***ϕ***/ℂ ratio. The general trend of penetration volume percentage is given by ***ϕ***/ℂ ratio.

### Droplet size and velocity distribution

As explained in the methods section, a model-cough is produced using a spray nozzle orifice by maintaining the droplet velocity reaching maximum value of 14 m/s, and droplet sizes ranging from 0-600 μm peaking around 60-70 μm, similar the characteristics of respiratory droplets ejected during a cough or mild sneeze as mentioned in the literature^9,28,36^. These droplets are shown to have significant number of smaller droplets in the aerosol range (< 100 μm), and the velocity scale is around 10 m/s. However, less number of larger size cough droplets (> 250 μm) contribute to more than 94% of the total volume. The Fig. 3 (a-g) shows the shadowgraphy images of the model cough impingement generated using a nozzle-orifice fitted inside the mouth of a manikin wearing different mask samples. The Fig. 3 (a) shows the model-cough spray coming out of the manikin’s mouth opening and it is clearly evident from the image that the spray consists of droplets having a wide range of sizes. The cone angle of the spray is shown in the figure as well (see Fig. 3 (a)). Since manikin head has been used, the spray coming out of the mouth exhibits a more realistic behavior by interacting with the manikin’s mouth edges leading to other effects like secondary breakup like phenomena.

**Fig. 3.**
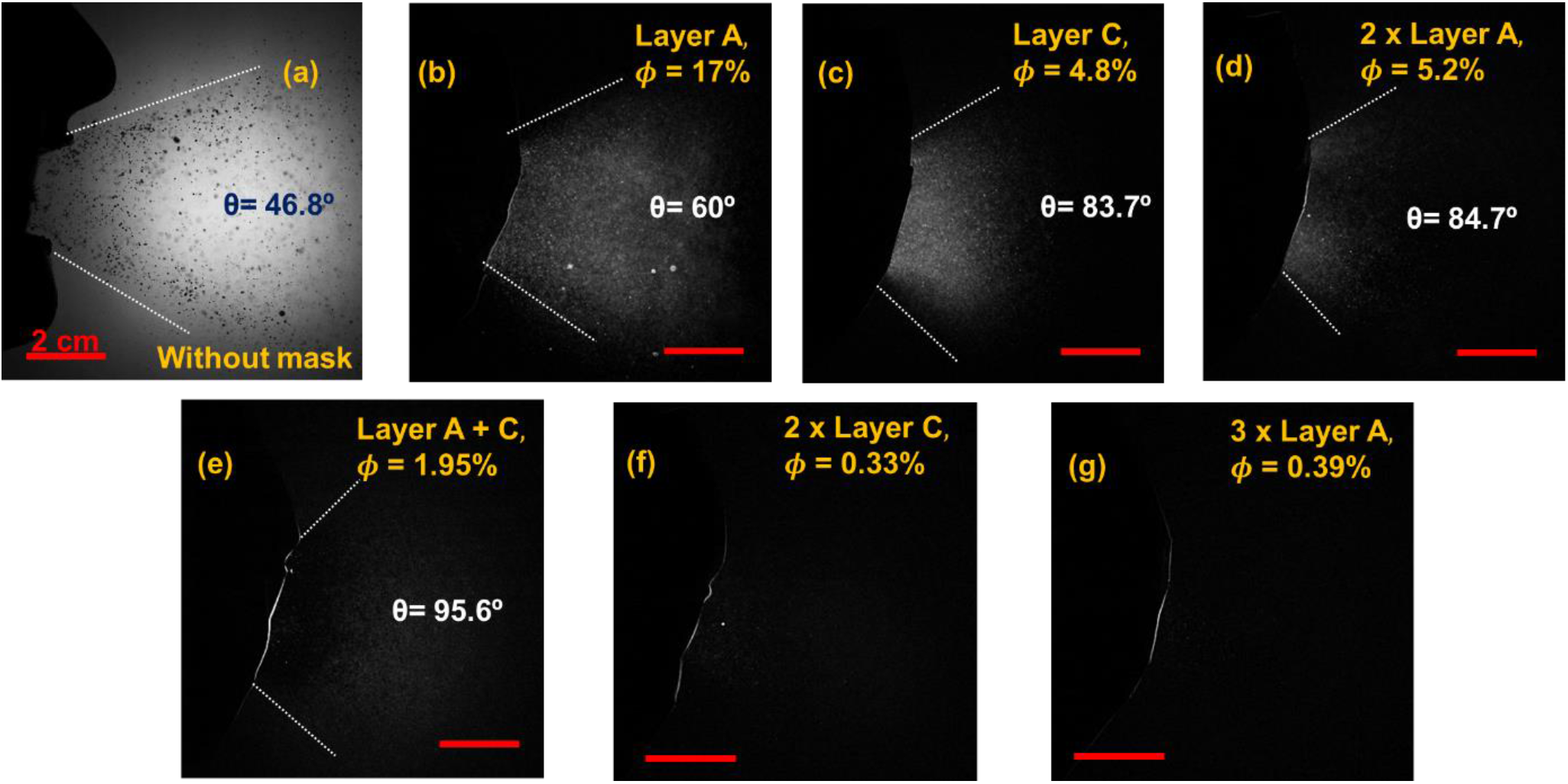
The figure shows the spray characteristics and the experimental images of spray impingement using a manikin with and without mask. **(a)** Shadowgraphy image of model-cough generated using spray nozzle in a manikin without mask, **(d-i)** Enhanced high-speed images of penetrated daughter droplets with different mask samples (in decreasing order of penetration volume percentage): **(d)** Layer **A, (e)** Layer **C, (f) 2 x** Layer **A, (g)** Layer **A + C, (h) 2 x** Layer **C, (i) 3 x** Layer **A**. The values of porosity (***ϕ***) for each sample, as well as the cone angle of the spray ejected into surroundings for the respective samples is shown. All the scale bars represent 2 cm.

Different test samples of masks are prepared and worn to the manikin head and model-cough impingement studies have been conducted. The penetrated daughter droplets in manikin experiments for the samples **A** and **C** have been shown in movie 1. The Fig. 3 (b-g) show the ejected daughter droplets after penetration through different type of masks (in decreasing order of the penetration volume percentage), (see table 4). The Layer **A** which has relatively larger pore size (**ϵ ∼ 45** μm) and porosity (***ϕ*** ∼ **17**%) results in considerable amount of large sized droplets along with aerosolization Fig. 3 (b). The samples, Layer **C** and **2 x** Layer **A** has similar pore size (**ϵ ∼ 30** μm) and porosity (***ϕ*** ∼ **5**%), however the latter exhibited significantly low penetrated droplets due to its higher fabric thickness compared to the former, as shown in Fig. 3 (c and d). It is also interesting to observe that the cone angle of the penetrated daughter droplet spray has increased to ∼ 60° while using Layer **A** and ∼ 80° for Layer **C** and **2 x** Layer **A** samples. This shows that the forward momentum of the impinging jet has been dissipated and distributed due to the presence of the mask. This effect is minimal in case of Layer **A** sample which exhibited relatively smaller cone angle due to its larger porosity resulting in relatively lower viscous dissipation. The Fig. 3 (e) corresponds to the sample which is the combination of Layer **A** and Layer **C**. It exhibited significantly reduced penetration volume compared to the **2 x** Layer **A** sample because of the presence of less porous Layer **C** (***ϕ*** ∼ **5**%) along with Layer **A** sample. The Fig. 3 (f and g) the two samples **2 x C** and **3 x A**, having very low porosity (***ϕ*** ∼ **0. 3**%) exhibit almost negligible amount of penetrated daughter droplets all of which are totally in the aerosolization range. The afore-mentioned model-cough generated in the experiments as shown in Fig. 3 replicates similar velocity and size distribution characteristics of an actual cough, as shown in Fig. 4 (b and c). The Fig. 4 (d-f) show the PDFs of the penetrated droplet velocity distribution for different samples which follow bell-shaped trend. The Fig. 4 (d and f) depicts the effect of addition of multiple layers to the incident spray. In both the cases, the velocity PDF curve is observed to shift towards left with the addition of each extra layer. This suggests that each additional layer contributes to the dissipation of initial kinetic energy of the impinging spray. It is also to be noted that the ratio ***t***_***m***_/***ϵ*** increases drastically from ∼10 to ∼53 with addition of two more layers in case of sample **A** (similarly for sample **C**, see table 4), suggesting increased viscous dissipation effects. This can be attributed to both increased viscous dissipation due to higher net fabric thickness and reduced effective porosity due to additional layers (see movie 2). The value of the peak is also observed to increase with additional layers suggesting more droplets get decelerated to lower values of velocity. The Fig. 4 (e) shows the effect of combination of mask layers with different porosity. The Layer **A** and Layer **C** individually have porosities ***ϕ*** ∼ 17% *and* 5%, allowing ∼ 16% *and* 4.5% of volume to penetrate through them respectively. The combination of these two layers shifts the curve to the left significantly because of the low porosity of the Layer **C** contributing to higher level of viscous dissipation additional to the effect of added fabric thickness (see movie 3).

**Fig. 4.**
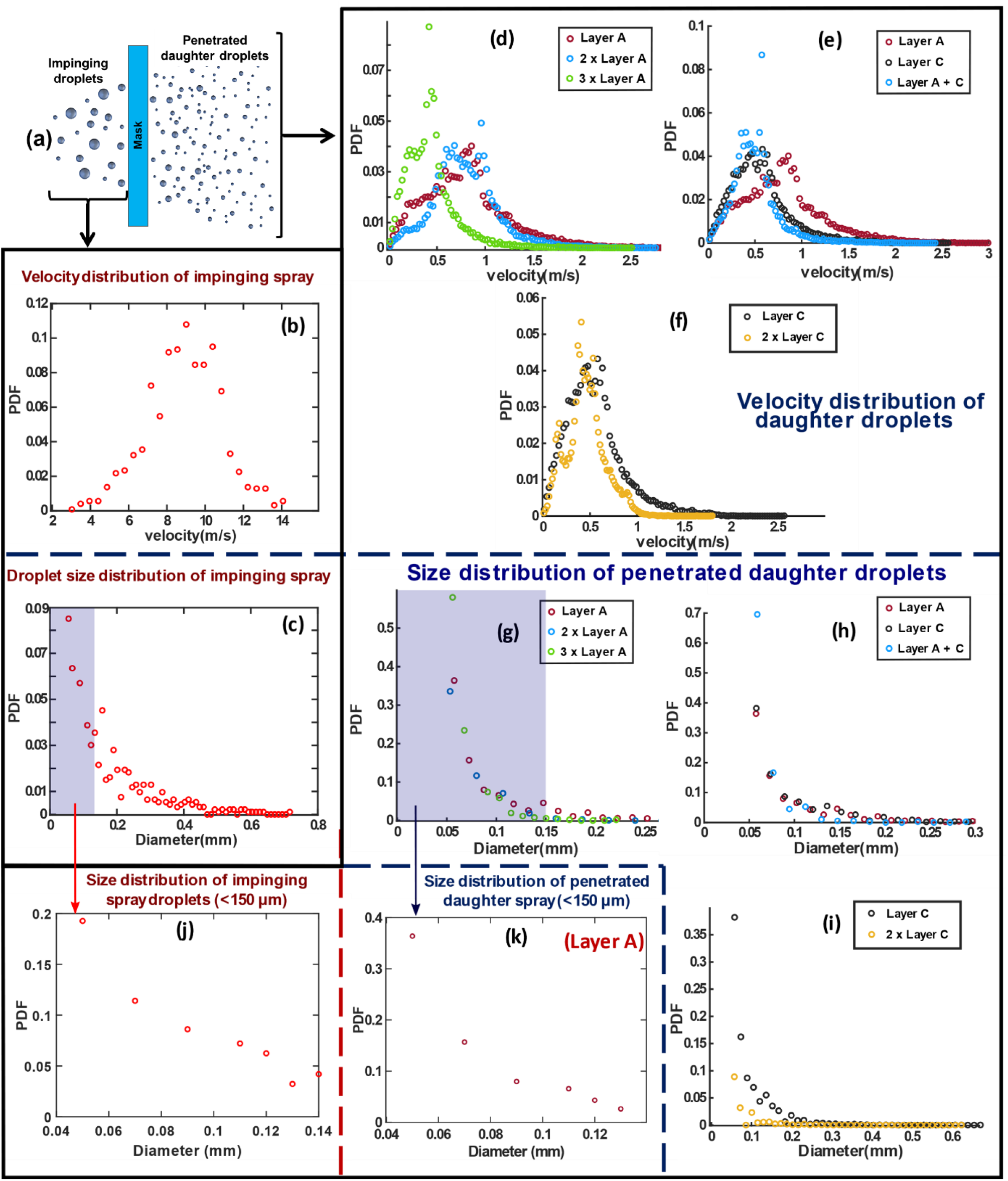
The figure shows the spray characteristics of spray impingement on mask surface. **(a)** The schematic depiction of spray impingement and penetration of daughter droplets through a mask, **(b)** Velocity distribution (PDF) for the impinging spray droplets, **(c)** Droplet diameter distribution (PDF) for the impinging spray, **(d-f)** Velocity distribution (PDF) for the penetrated daughter droplets for different samples: **(d)** Layer **A, 2 x** Layer **A**, and **3 x** Layer **A**, showing effect of number of layers, **(e)** Layer **A**, Layer **C**, and Layer **A + C**, showing the effect of combination of different samples, **(f)** Layer **C**, and **2 x** Layer **C, (g-i)** Droplet size distribution (PDF) for the penetrated daughter droplets for different samples: **(g)** Layer **A, 2 x** Layer **A**, and **3 x** Layer **A, (h)** Layer **A**, Layer **C**, and Layer **A + C, (i)** Layer **C**, and **2 x** Layer **C, (j-k)** Droplet size distribution (PDF) in aerosolization range (< 150 μm): **(j)** For the impinging droplets on the mask, **(k)** For the penetrated daughter droplets for sample **A**.

In all cases the velocity range of penetrated droplets is in the range of 0-2 m/s, suggesting a physical obstruction like a mask can decelerate the high velocity (∼ **10** − **14 *m***/***s***) cough droplets to the order of < 2 m/s. However, having better mask properties such as lower porosity and pore size or higher thickness leads to a higher proportion of droplets that are decelerated to lower velocities (< 1 *m*/*s*). This extent of deceleration of the impinging cough droplets is essential to contain the spread of these pathogen loaded droplets from an infected person. However, the droplets in aerosolization range can remain suspended in air for longer durations and can be transferred to different locations by the air flow which is another important issue.

The Fig. 4 (g-i) show the PDFs of the droplet size distribution after penetration through the corresponding mask samples. The diameter of the penetrated daughter droplets depends on the ligament size and velocity, which is governed by the mask pore size (***ϵ***), fabric thickness (***t***_***m***_)^21^. The pore size contributes to capillary effects and viscous dissipation whereas fabric thickness affects only viscous dissipation. In case of Fig. 4 (g), addition of extra layer for the sample Layer **A** leads to decrease in effective pore size and increase in fabric thickness. The increase in fabric thickness reduces the ligament velocity and shorter ligaments due to viscous dissipation. Additionally, the decrease in pore size leads to the formation of smaller daughter droplets after ligament breakup. This is reflected in the Fig. 4 (g), where the probability decreases for the formation of larger droplets and increases for the formation of smaller droplets due to the additional layer (see movie 2). Similar behavior is observed in case of Layer **C**, (see Fig. 4 (i)). Additionally, curve shifts significantly to the left in case of layer **C**, which can be attributed to the lower pore size and higher thickness of Layer **C**. In case of Fig. 4 (h), the similar trend is observed where with the combination of the samples **A** and **C**, the PDF curve shifts to the left and height of the curve which is present in the lower diameter range increases. Hence, the probability of the formation of smaller droplets increases. The size distribution has been plotted for droplets belonging to the aerosolization range (< 150 μm) for impinging droplets and penetrated droplets in Fig. 4 (j,k) respectively. These plots give insight into the aerosolization occurring due to ligament breakup of smaller droplets. The probability of smaller sized droplets is observed to be higher for the daughter droplets after penetration in aerosolization range (see Fig. 4 (k)). This is because even smaller droplets contribute to aerosolization, as shown in Fig. 2 (c).

It is to be noted that in all the droplet size distribution PDFs shown, due to the experimental limitations involved in imaging the spray, the minimum droplet diameter that can be measured with decent accuracy is 50 ± 23.8 μm. Hence, the droplet diameter PDFs does not have any data points below diameter value of 0.05 mm which forms the left portion of the bell-curve trend for penetrated daughter droplet diameter PDF. The PDFs of the samples Layer **B**, **3 x** Layer **C** are not plotted as the penetration is not detected in these samples.

Comparing the two samples Layer **A** and Layer **C**, the pore size of sample **A** is larger compared to sample **C**, which tends to the formation of thicker ligaments in the sample **A** leading to larger sized droplets (see supplementary fig. S2 (a) and fig. S2 (d)). However, the fabric thickness of sample **A** is significantly lower which tends to increase the ligament velocity and hence enhance ligament breakup phenomenon. Additionally, the PDFs of sample **A** and sample **C** look similar with similar type of droplet distribution having peak at around 60-70 μm. Hence, it can be inferred that these two effects of pore size and fabric thickness counteract each other in influencing the droplet size distribution in these two samples, comparatively (see Fig. 4 (g and i)). This effect is also evident from the fact that the highest probable velocity is higher for sample **A** compared to sample **C** which can be attributed to lower loss of initial kinetic energy to the viscous dissipation and lower thickness (see Fig. 4 (d and f)). Even though Layer **C** has a significantly lesser number of daughter droplets due to its low porosity, the penetrated daughter droplets have similar proportion of different ranges of droplet sizes in both the cases **A** and **C**, as shown in the experimental shadowgraphy images (see supplementary fig. S2 (a) and fig. S2 (d)).

Furthermore, it is interesting to note that in case of samples Layer **A, 2 x A**, a very small number of larger size droplets (> 150 μm) are also present along with the dominant 0-100 μm range daughter droplets (as shown in supplementary fig. S2 (a,b)). This shows a mild possibility of the presence of larger droplets even in case of double layers when the layers have high porosity similar to Layer A (***ϕ*** ∼17%). However, this effect was not significant in case of Layer **C** either single or double layer of sample **C**, due to its smaller pore size as well as larger fabric thickness, as shown in Fig. 4 (f) and supplementary fig. S2 (d, f). When performance of the two samples, **3 x** Layer **A** and **2 x** Layer **C** are compared, the higher fabric thickness in **3 x A** contributes to the lower penetration volume due to the dominant viscous dissipation. However, the penetrated daughter droplets in case of sample **3 x A** have significant number of relatively larger droplets (100-200 μm) even though it is a three-layered configuration, compared to the two-layered configuration of **2 x C** which is having lower porosity (see Fig. 4 (g,i)). Comparing the two samples **A + C** and **2 x C**, it is evident that even though the penetration volume percentage is similar (∼0.35 %) in both the cases, the sample **A + C** exhibited the presence of relatively larger sized penetrated droplets (> 100 μm) compared to the sample **2 x C** (Fig. 4 (h,i) and supplementary fig. S2 (e,f)). This is attributed to the larger pore size of the layer **A**. The spray impingement phenomena in case of sample **2 x A, A + C, 2 x C** are shown in movie 4.

Another observation from the shadowgraphy images is that the lesser porous samples exhibit significant amount of bounced back volume which is retained on top of the mask without penetration. From these experiments, it can be inferred that the lower effective porosity of the mask due to the additional number of layers leading to higher fabric thickness can lead to significantly lower volume penetration compared to the samples having similar or relatively lower pore size. This is because of the dominance of the viscous dissipation effects which resist the penetration of the droplets in case of masks having higher ***t***_***m***_/***ϵ*** ratio.

The ***ϕ***/ℂ ratio is effective in incorporating the effect of global parameter porosity (***ϕ***) on spray impingement in addition to the pore level criteria based on the local parameters like pore size (***ϵ***) and fabric thickness (***t***_***m***_). Additionally, the ***ϕ***/ℂ ratio is a good tool to estimate the extent of penetration of a spray through a given mask fabric. It is to be noted that if the pore sizes are varying drastically between the samples, the effect of ***ϕ*** on penetration will not be straightforward, as the pore size (***ϵ***) can affect both viscous dissipation as well as capillary effects. Lower pore size is needed for higher viscous dissipation and larger capillary forces that resist volume penetration. This leads to lower penetrated volume but significant number of smaller daughter droplets in aerosolization range will be formed. However, low porosity and relatively higher pore size is also another option to consider for a mask as shown by Krishan et al., where lower net volume penetration can be achieved with lesser extent of aerosolization effect^21^. In addition to all these intercoupled effects, usage of higher thickness material for mask layer is beneficial in reducing penetration volume. However, minimum three layers are necessary in order to reduce the effect of aerosolization which can minimize the potentially harmful outbreak of contagious diseases.

The current experiments try to replicate the droplet sizes and velocities involved in a coughing event using a spray, to study the effects of mask properties on the droplet penetration. It is evident from the data that the droplets of different sizes (d > ***ϵ***) atomize into even smaller daughter droplets, resulting in significant aerosolization. This suggests that the penetration criteria are also valid for smaller droplet range (d < 100 *μm*) which contribute significantly to the aerosolized penetrated droplets (see Fig. 4 (j,k)). It is to be noted that, the scope of the current experiments is only limited to the droplet dynamics of the spray impingement and its penetration through the mask. However, the effect of air flow and turbulence involved during an actual coughing event will result in the transportation of these aerosolized ejected droplets^34,29^. This presence of air flow and turbulence increases the range of infection compared to the distances associated with the velocity of these individual penetrated droplets. Hence, the respiratory jet and turbulence effects have to be superimposed over the velocity scales of the penetrated droplets ejected, while considering the range of infection around an infected person wearing a mask.

### Effect of mask usage by secondary manikin during model cough of primary manikin

From the experiments, the penetrated droplets through the mask fabric are shown to be in lower diameter regime with significant number of droplets in the aerosolization range. The velocity scale of the penetrated daughter droplets is in the range of 0-2 m/s. This section discusses the fate of ejected droplets through a mask, when a second manikin is placed in the path of their trajectory. Experiments have been conducted to replicate the situation similar to social conversations and to determine the effectiveness of the mask usage.

The two manikins are placed at a distance of 10 cm from each other (see Fig. 5 (a,b)). The primary manikin (representing the infected person wearing mask) has been used for inducing model-cough, generated using a spray of DI water. A dark colored dye is added to the DI water for detection of penetrated droplets. On the receiving end, the mouth portion of the secondary manikin (representing susceptible person) has been covered by a white cloth to absorb any droplets impinging on it, and mask is worn on top of it, as shown in Fig. 5 (b). Experiments were conducted as explained in methods section (see Fig. 7), using the susceptible person manikin both with and without the mask, during the cough event from infected person manikin. To study the penetration specifically, all these experiments were performed using high porosity single layer mask (**Layer A, *ϕ*** ∼ 17%).

**Fig. 5.**
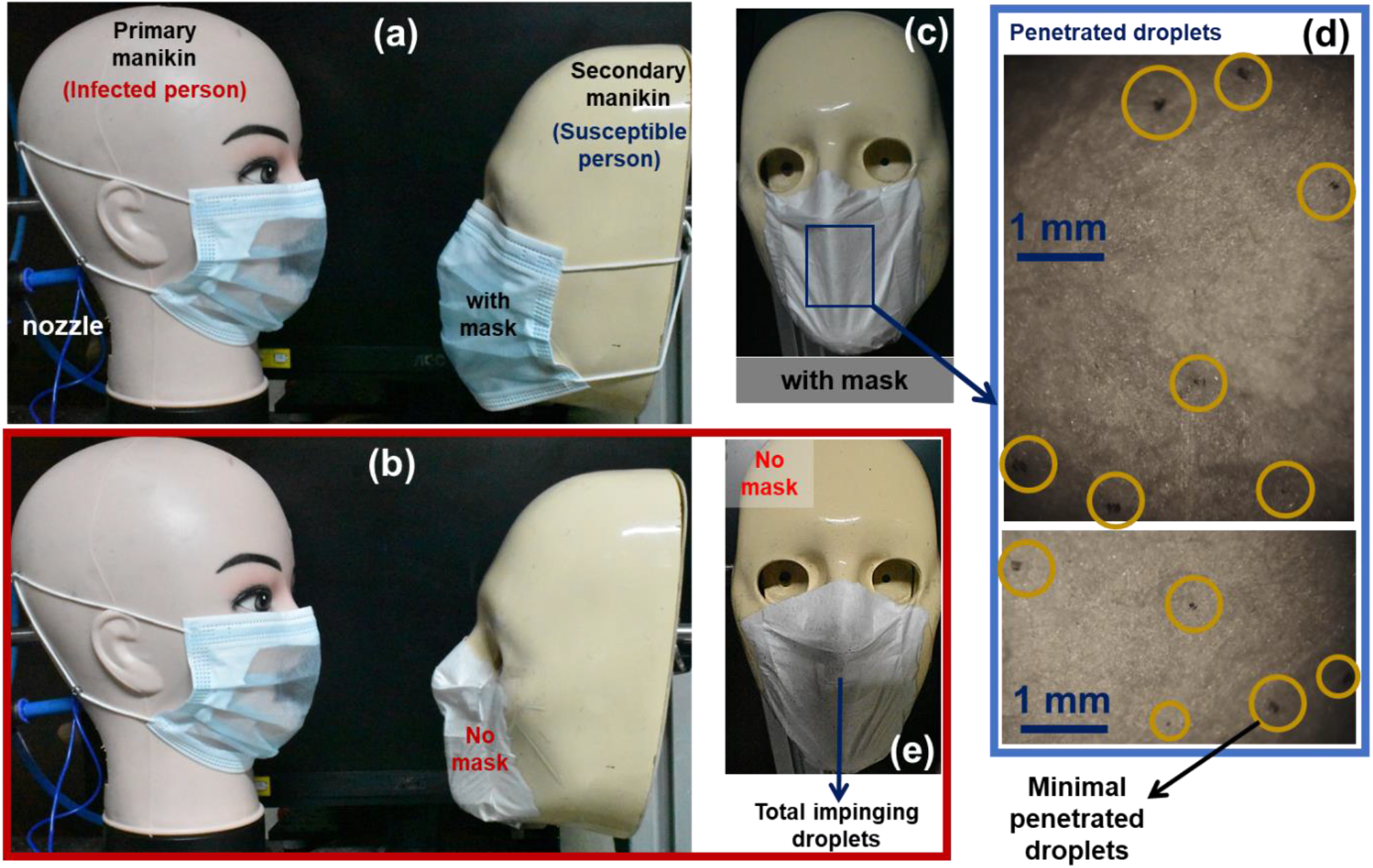
The figure shows double manikin experiments to investigate the fate of penetrated droplets from primary manikin (representing infected person with mask) in the presence of secondary manikin (representing susceptible person) in the vicinity. **(a)** two manikins placed at 10 cm distance with both wearing a mask, **(b)** two manikins facing each other with secondary manikin not wearing mask. **(c)** Snapshot of penetrated droplets on secondary manikin wearing a mask against repeated cough cycles, **(d)** the zoomed-in microscopic images of the penetrated droplets on the susceptible person manikin wearing mask, showing minimal penetration, **(e)** Snapshot of secondary manikin without wearing mask, showing total amount of impinging droplets ejected from the infected person manikin. All scale bars are shown.

When the mask is worn on the susceptible person, repeated cough events of the primary manikin (infected person) led to penetration (see Fig. 5 (c,d)). This is evident from the small number of penetrated droplets detected on the white cloth pasted on the mouth of susceptible person (see Fig. 5 (d)), when compared to total number of droplets without mask, as shown in Fig. 5 (e). The minimal penetration through the mask of the receiving manikin is because of the reduced velocity scales (0-2 m/s) of the ejected droplets, as they pass through the mask of infected person^22^. However, these smaller number of droplets potentially can cause infection when they penetrate the mask of susceptible person. This shows that it is not sufficient to reduce the risk of infection even if the single layer masks are worn by both infected and susceptible person. Thus, the experiments clearly suggest the necessity of the need of multiple layers as well as social distancing, in mitigating the spread of contagious respiratory diseases.

### Penetration dynamics of virus-emulating nanoparticle laden surrogate respiratory droplets

From the experiments, it has been shown that the mask blocks the significant volume of cough droplets ejected out. Nevertheless, the effectiveness of mask to block the virions present in infected respiratory droplets also needs to be investigated. Different particle loadings of 100 nm polystyrene nanoparticles (R-100) in the mucin solution (0.9% by wt. NaCl, 0.3% by wt. gastric mucin, 0.05% by wt. DPPC added to DI water), has been used to replicate the virion laden surrogate respiratory droplets, to investigate the mask filtration. These solutions mimic the fluid dynamics of virion-laden droplets, although they do not have the mechanical or chemical properties of virions^6^. The virus-emulating nanoparticle laden droplets are then impinged onto the face mask, and the deposition on the mask surface as well as penetrated nanoparticles has been identified from the fluorescence microscopy images. The fluorescence microscopy images have been superimposed on the bright-field microscopy images to show the corresponding location of the nanoparticle entrapment, on the mask layer.

The impinging spray generated from the spray orifice, using the nanoparticle laden mucin solution at different particle loadings has been found to exhibit similar droplet size distribution as that of DI water (see Fig. 6 (a)). Fig. 6 (b) shows the schematic depiction of the virus-emulating nanoparticle laden droplet spray penetration, which shows the entrapment as well as penetration of virions through the mask. The fluorescence microscopy images of nanoparticles present in the penetrated droplets has been depicted in Fig. 6 (d-f). The size distribution of the penetrated droplets is observed to remain similar for the viral-loaded surrogate respiratory droplets as well (see Fig. 6 (c, d-f)). The Fig. 6 (g-i) shows the overlaid bright-field and fluorescence microscopy images on the impinged side of the mask, indicating the presence of trapped nanoparticles. This nanoparticle deposition on the mask indicate the entrapment of virions. This mandates the necessity to follow proper disposal methods for handling face masks after utilization. Experiments show that the entrapment as well as penetration of virus-emulating nanoparticles increase with increase in initial loading rates (see Fig. 6 (d-f, g-i)). It is further observed that there is a marginal increase in the tendency for the formation of larger sized droplets (∼400 *μm*) at very high particle loading (10^9^ particles/ml). It is evident from Fig. 6 (g-i) that the masks block and entrap a significant amount of nanoparticles (virions) along with the droplet volume, during the penetration. The virus-emulated nanoparticles were observed to be entrapped in the crevices and at the junction of the fibres on the mask layer. This suggests that the lower porosity of the mask contributes to the higher availability of the morphological surfaces on the mask which are capable to entrap the virions. Thus, the lower porosity results not only in net reduction in the percentage volume-penetrated through the mask, but also provides more surface area for the entrappement of nanoparticles. This further suggests that the morphology of the mask comprising of different type of crevices can contribute to the virions entrappement.

**Fig. 6.**
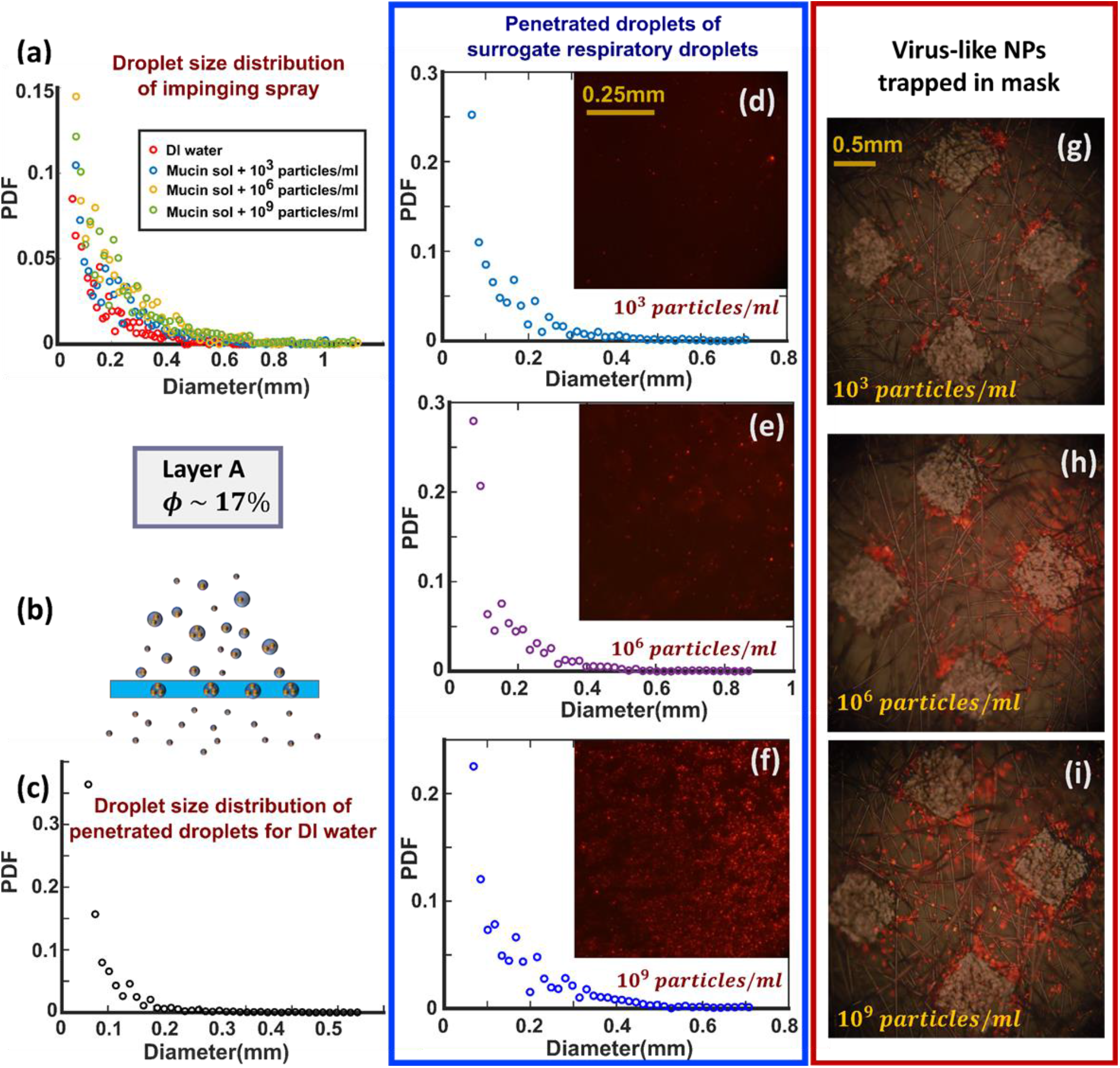
**(a)** The figure shows the impinging spray characteristics to be similar for the nanoparticle (NP) laden mucin solution for different particle loading rates and DI water. **(b)** The schematic of the virus-like nanoparticle laden spray droplets penetration has been shown. **(c)** Figure shows the droplet size distribution of penetrated droplets through **Layer A** for DI water. **(d-f)** Figures show the droplet size distribution (PDF) of penetrated droplets through **Layer A** using: **(d)** 10^3^ NP particles per ml of mucin solution, **(e)** 10^6^ NP particles per ml of mucin solution, **(f)** 10^9^ NP particles per ml of mucin solution. **(d-f)** The fluorescence microscopy images of virus-like nanoparticles corresponding to the penetrated droplets has also been shown as subfigures for the respective solutions. **(g-i)** Figure shows the virus-like nanoparticles (NP) trapped in the mask for particle loadings: **(g)** 10^3^ NP particles per ml of mucin solution, **(h)** 10^6^ NP particles per ml of mucin solution, **(e)** 10^9^ NP particles per ml of mucin solution. **(g-i)** Figure shows the overlaid bright-field and fluorescence microscopy images indicating the presence of fluorescent virus-like nanoparticles embedded in the mask. All scale bars are shown.

## Conclusion

The droplet penetration phenomenon through a mask is dependent on both surface tension as well as viscous dissipation. Both the criteria are to be considered together to determine the possibility of penetration. However, the relative importance of one criterion over the other is dependent on the type of the mask and its properties like pore size and fabric thickness. In the current experiments it is found that the viscous dissipation criterion is more suitable for the samples with small pore size. However, these two criteria focus on the pore level dynamics only and do not account for the penetration of the spray. Hence, a new parameter has been formulated based on the area available for the spray and spray density to estimate the possibility of penetration. This parameter (***ϕ***/ℂ ratio) offers a new penetration criterion in addition to the two-step criteria, to incorporate the global effects of properties like porosity (***ϕ***) and spray density. This parameter can also give a qualitative estimate of the probability of penetration and extent of penetrated volume in case of a spray. Both the two-step criteria as well as the ***ϕ***/ℂ ratio criteria are necessary to predict the penetration of a spray accurately. This parameter can give a general trend of the percentage volume-penetrated, however effect of the local parameters should also be considered for accurate estimation.

To get insights into the aerosolization during penetration, the effect of mask properties on the penetrated droplet size as well as the velocity distribution has been investigated. The lower pore size results in higher probability of the formation of droplets in the aerosolization range and lower velocity scales. The higher fabric thickness increases viscous dissipation, resulting in higher probability for the ejected droplets to have lower velocity. This is also reflected in case of using additional layers due to the reduction of effective pore size as well as the increased effective fabric thickness. The effect of pore size and combination of different mask layers has been investigated in the study. The multilayer mask will result in significant reduction in the volume of the penetrated daughter droplets which will reduce the amount of pathogen loading released into the surroundings. However, reduction of aerosolized droplets along with maintaining low penetration volume percentage is ideal scenario, as small droplets can be suspended for longer durations increasing the infection potential. The current investigation gave insights into this by having a multilayer mask having combination of different layers with relatively high pore size as well as low pore size. It is to be noted that this will only be effective if the combination can result in low penetration volume percentage. However, further investigation is necessary to get better insight into the effect of pore size.

The penetration characteristics of the spray show that all ranges of droplet diameters (d > ***ϵ***) contribute to aerosolization. The smaller droplets (d < 100 *μm*) tend to atomize into even smaller sized droplets resulting in higher degree of atomization along with larger droplets (d > 250 *μm*). This aerosolization effect, which is purely based on droplet dynamics, coupled with the turbulent flow associated with respiratory jets during the cough will potentially increase the infection potential to larger distances by the transportation of aerosolized droplets. Hence, all the penetration criteria along with the turbulence effects are to be considered in estimating the potential infection zone around an infected person. It has been observed from the manikin studies that penetration is observed at the receiving end, even when both the infected as well as susceptible person wear single layer mask. This suggests that both social distancing as well as multilayered mask are necessary to efficiently block the viral loaded respiratory droplets from spreading the infection. Additionally, experiments show that the masks block the impinging droplets along with the virus-emulating nanoparticles. The data also suggests that the virus-emulating nanoparticles are entrapped in the morphological crevices present on the mask.

## Methods

Experimental investigation has been conducted to study the efficacy and penetration criteria for a model-cough (spray) impingement on mask surface using high speed shadowgraphy imaging. The cough or sneeze event simulated with DI water spray. The spray is generated using a pressurized liquid chamber, a nozzle orifice and a solenoid valve. Arduino code is used to actuate the solenoid valve for a specific opening time. When triggered, the solenoid valve opens and closes based on the preset time delay, allowing the pressurized DI water to eject out of the nozzle in the form of a spray. It must be noted that the respiratory droplet velocity and size vary depending on the specific event: normal breathing, coughing, sneezing etc. In this study, the velocity scale and droplet sizing of the incident spray is maintained in the range corresponding to respiratory droplets involved in actual cough or mild sneezing events, as indicated in the literature. In current experiments, the droplet velocity range is around 10 m/s with maximum velocity reaching up to 14 m/s and the droplet diameters distribution range of 0-600 μm, peaking around 60-70 μm (similar range as shown by Duguid^9^) (see supplementary fig. S1). Additionally, the spray cone size at the point of impact on the mask is set to be in the range of ∼3-4 cm, as shown for typical cough droplets^33,34^.

The global characteristics of spray impingement through a mask has been investigated using a manikin head with the afore-mentioned nozzle orifice used as the source of the spray from the mouth opening, (see Fig. 7). Effort has been made to replicate the spray similar to that of an actual cough or sneeze as explained before by matching the parameters like droplet size, velocity range, and spray size. The main objective is to check for the penetration characteristics, and get comprehensive insights into the spray impingement and penetration phenomena. An actual spray is used for impingement in current study instead of single droplet, which has been investigated in our previous work. Using a manikin, the spray penetration characteristics has been investigated qualitatively. The physical proximity and dimensions with the manikin and spray are maintained similar to that of an actual human cough, with and without wearing a mask.

**Fig. 7.**
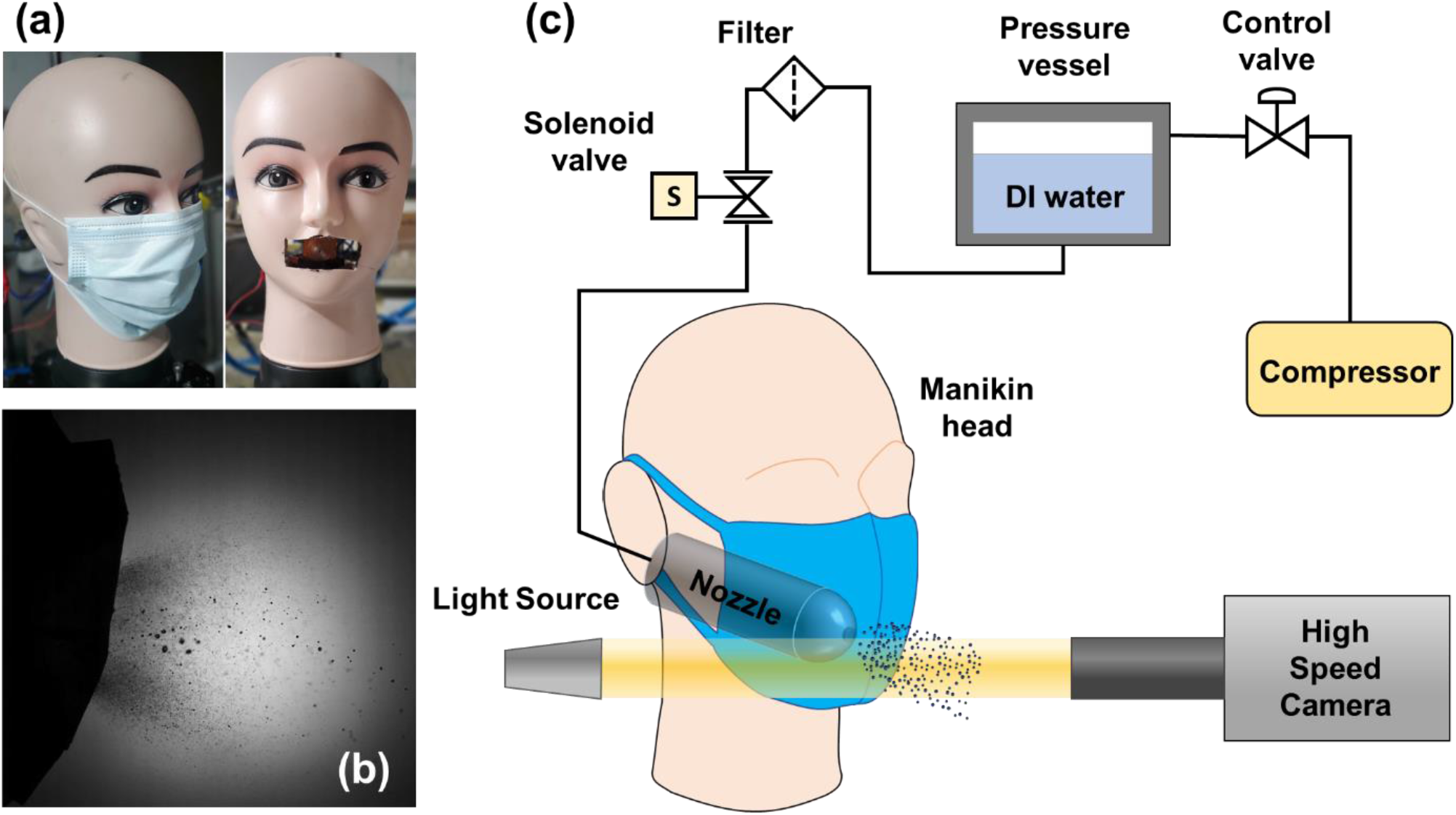
**(a)** Manikin with and without mask. Spray nozzle orifice is shown on the right side. **(b)** Shadowgraphy image showing the daughter droplets after penetration for Layer A. **(c)** Manikin experimental setup: High-speed cameras were used to capture the side view of the Manikin experiments for different mask samples. Pressurized DI water is connected to the nozzle through a solenoid valve.

Additionally, a local investigation has been conducted to check the spray impingement phenomenon quantitatively on the mask layers which are mounted on a post. A more fundamental approach has been taken to study the correlation between mask morphological characteristics and penetration characteristics like the size and velocity distribution of the penetrated daughter droplets. The effect of the presence of multiple layers on the spray impingement has also been investigated in detail with different combinations of mask samples. DI water has been used to mimic the respiratory droplet spray ejected onto the inner side of the mask from an infected person (see Fig. 7 and 2). The surface tension of DI water (*σ* = 72 mN/m) was found to be similar to that of cough droplets (*σ* = 65.9 mN/m)^22,40^. However, the other fluid properties may vary between the model-cough droplets used and the actual cough droplets. Nevertheless, in the scope of current study of spray droplet penetration, only surface tension, viscosity and viscoelasticity are relevant fluid properties. Furthermore, it has been shown that the effect of the droplet fluid properties on penetration becomes less critical at high impact velocities (∼ 10 m/s)^41^. Additional experiments with surrogate respiratory liquid (0.9% by wt. NaCl, 0.3% by wt. gastric mucin, 0.05% by wt. DPPC added to DI water, i.e., mucin solution)^22,42^ have been conducted. It showed no noticeable difference in the penetration characteristics, suggesting an insignificant influence of droplet fluid properties at high impact velocities. Hence, DI water has been used for the spray impingement experiments. In addition to that, experiments have been performed using the virus-emulated nanoparticle laden surrogate respiratory liquid (mucin solution), to generate a model-cough spray for impingement^5,22^. The 100 nm sized R-100 polystyrene nanoparticles were used as model material for the viral loading based on similarity in geometrical properties (although they differ in biological and chemical properties)^6^. Further, the filtration of viral loading by the mask has been investigated using the nanoparticle laden surrogate respiratory liquid at different particle loading. Experiments have been conducted further, by placing a second manikin (representing a susceptible person with and without the mask) in the path of penetrated droplets ejected from the primary manikin (infected person), to check the effect of mask usage by the susceptible person.

### High-Speed Shadowgraphy

A high-speed camera (Photron SA5) coupled with TOKINA MACRO 100 F2.8 with 36 mm extension tube (for side view) has been used to focus on the plane of descent of the spray (see Fig. 8). The side view of the spray impingement has been captured using a Mercury lamp light source as backlighting to study the penetration dynamics. The camera acquisition rate was kept at 10000 frames per second with shutter speed of 93000 cycles per second for the shadowgraphy imaging, at a resolution of 896 × 848 as shown in Fig. 8. Due to experimental limitations, uncertainty in measurement of droplet size is minimized by using adaptive thresholding to avoid the highly out-of-focus stray droplets. The droplets size and velocity were determined using object detection on the shadowgraphy images by binarizing the raw images with adaptive thresholding. From this, the line-of-sight-area of the droplets can be obtained from which the equivalent diameter is calculated to estimate the volume of each droplet. The data processing has been performed for 20 experimental runs for each case in order to obtain the probability distribution function of the droplet size as well as the droplet velocity. The uncertainty in droplet diameter measurement is ± 23.8 μm and velocity measurement is ± 0.12 m/s. It is to be noted that only those set of frames were considered for data measurements in which the spray inlet distribution is similar to that in the literature for actual cough droplets^9^.

**Fig. 8.**
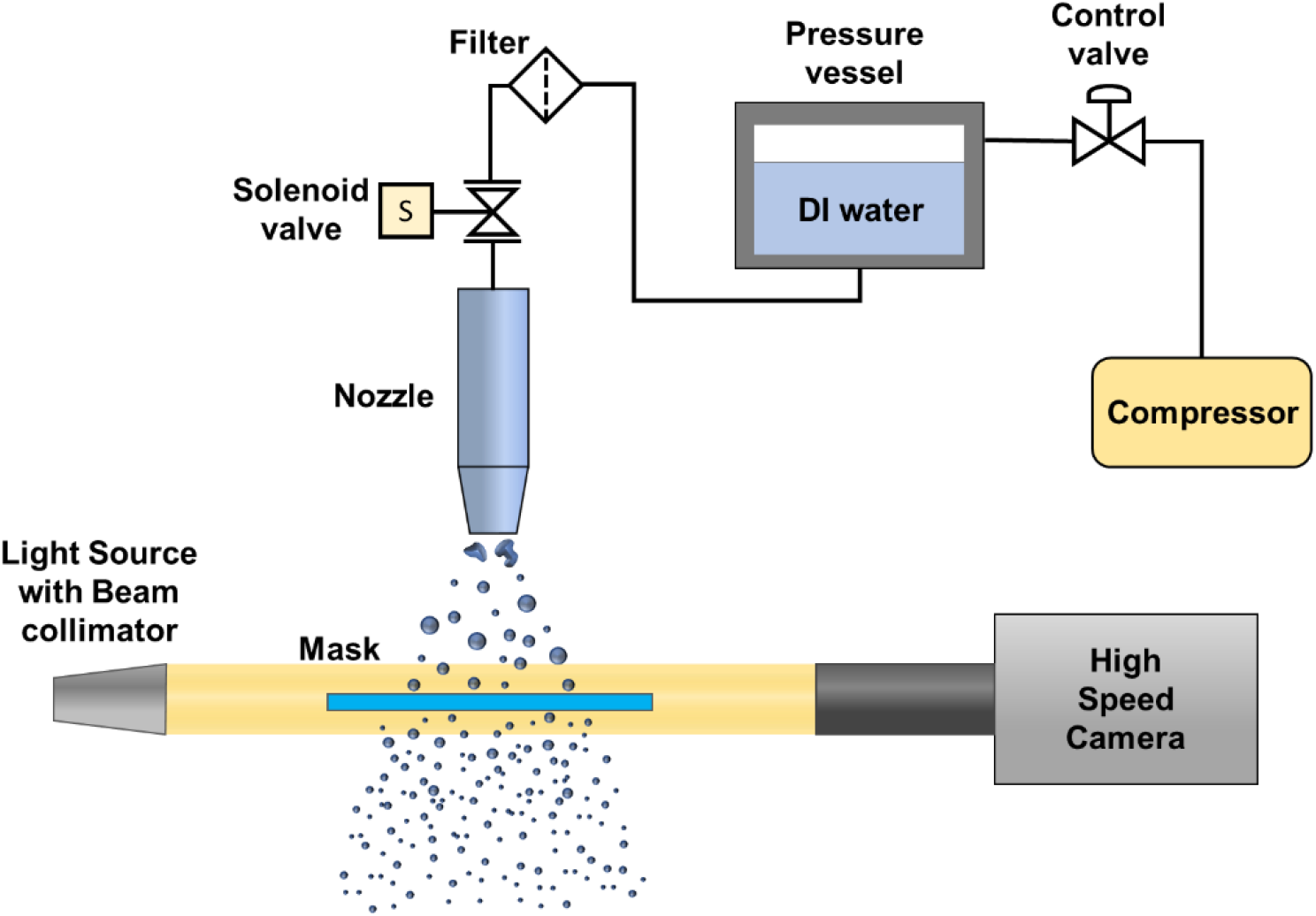
The experimental setup used for the fundamental investigation: High-speed cameras were used to capture the side view of the spray impingement on the surface of different mask samples. A high intensity light source has been used to capture the high-speed dynamics of spray impact.

### Mask sample characterization

In the current study, different layers (referred to as layer A, B, C) used in commercially available standard surgical masks have been considered as test samples which have varied range of properties like porosity (***ϕ***), fabric thickness (*t*_*m*_) and pore size (***ϵ***). Experimental investigation has been conducted using different combinations of these samples in single, double and triple layer configurations. The samples were characterized based on the afore-mentioned properties like pore-size and porosity which were calculated from the magnified cross-sectional shadowgraphy images of the samples, as shown in Fig. 9. Thresolding and binarization has been performed on these images to get the net area of the empty gaps or spaces available in the fabric sample (*A*_*pores*_).

**Fig. 9:**
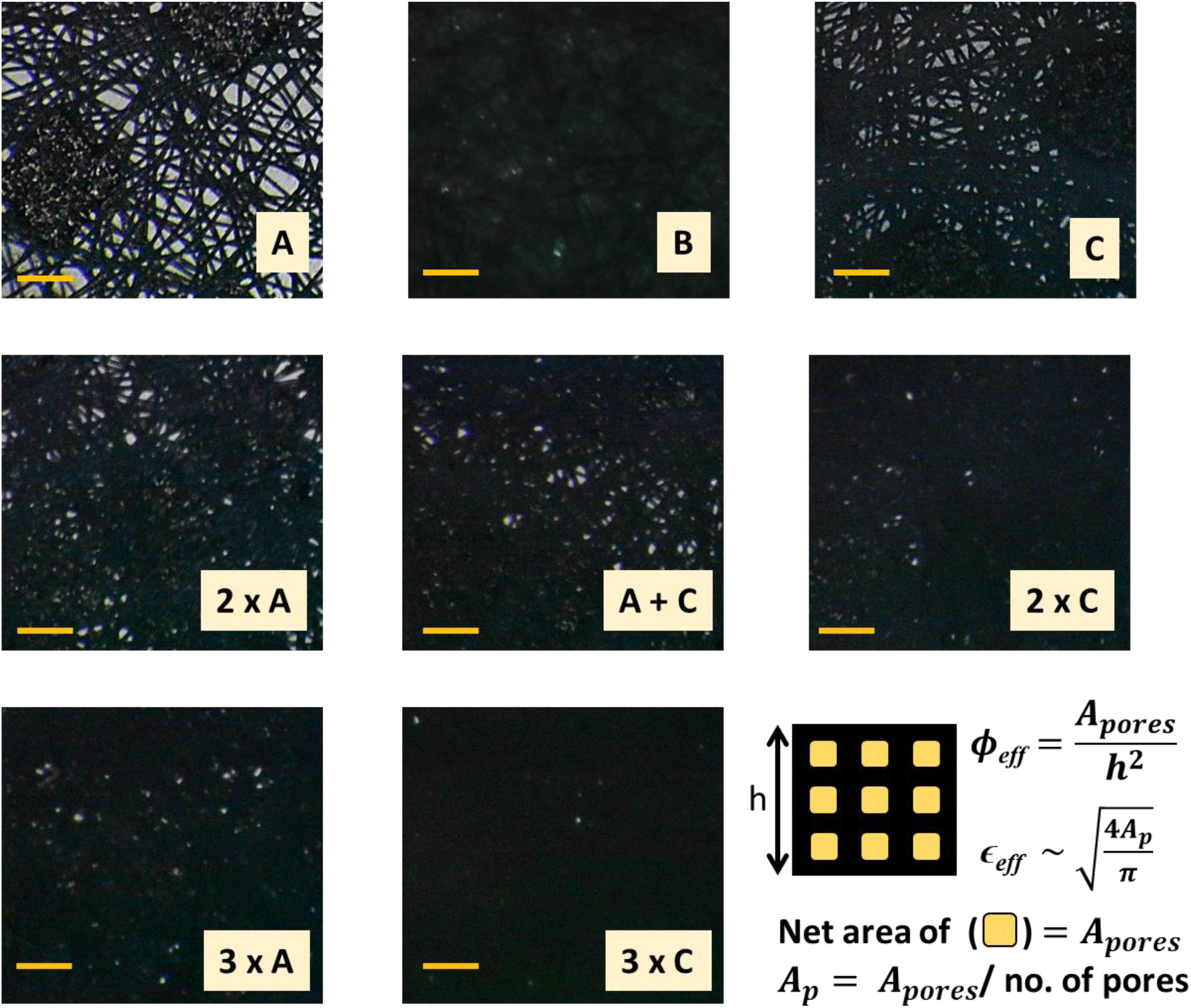
Magnified cross-sectional images (shadowgraphy) of each of the test samples as tabulated in table 1. The samples are: Layers **A, B, C** (single layer samples), 2 x **A, A** + **C**, 2 x **C** (double layer samples) and 3 x **A**, 3 x **C** (triple layer samples). All scale bars represent 300 μm. The methodology for obtaining pore size and porosity is shown in bottom-right corner.

The ratio of the net pore area obtained (*A*_*pores*_) and the total area of the fabric sample under consideration (*h*^2^) gives the value of porosity (***ϕ***) for the corresponding sample. The pore-size (***ϵ***) is obtained from the individual pore area *A*_*p*_ which is the ratio of *A*_*pores*_ and total number of pores present. With the addition of an extra layer, the amount of gap or pores available in the sample for the droplets to pass will decrease. Hence, effective value of porosity and pore-size are considered using the similar method for multilayered samples as well. The different test cases considered in this study with the sample properties in single, double and triple layer configurations are tabulated in the table 1. It is to be noted that, the multilayer configuration of layer **B** has not been considered, because the layer **B** alone has very low value of porosity which leads to negligible porosity when used in multilayer configuration.

## Data Availability

All data produced in the present work are contained in the manuscript

## Data Availability

The data is available in the manuscript and additional data is provided in the supplementary material.

