## Supplementary file for "Insights into spray impingement on mask surface: effect of mask properties on penetration and aerosolization of cough droplets"

**Nomenclature**

| $A_{mask}$ | Area of mask | $\boldsymbol{E}_{\boldsymbol{d}}$ | Viscous dissipation |
| --- | --- | --- | --- |
| $A_{p}$ | Individual pore area | h | Dimension of fabric sample under consideration |
| $A_{pores}$ | Net pore area | Re | Reynolds number |
| $A_{spray c/s}$ | Net cross-sectional projected area of spray droplets | $t_{m}$ | Fabric thickness |
| d | Droplet diameter | V | Impact Velocity |
| $\boldsymbol{E}_{\boldsymbol{k}}$ | Kinetic Energy | We | Weber number |

**Symbols**

| $\sigma$ | Surface Tension | ρ | Liquid density |
| --- | --- | --- | --- |
| $\boldsymbol{\phi}$ | Porosity | $\mu$ | Dynamics viscosity |
| ***ϵ*** | Pore size | $\mathbb{C}$ | Cross-sectional Spray areal density |

**Supplementary file includes:**

**Figs. S1-S2**

**Movies-S1, S2, S3, S4**

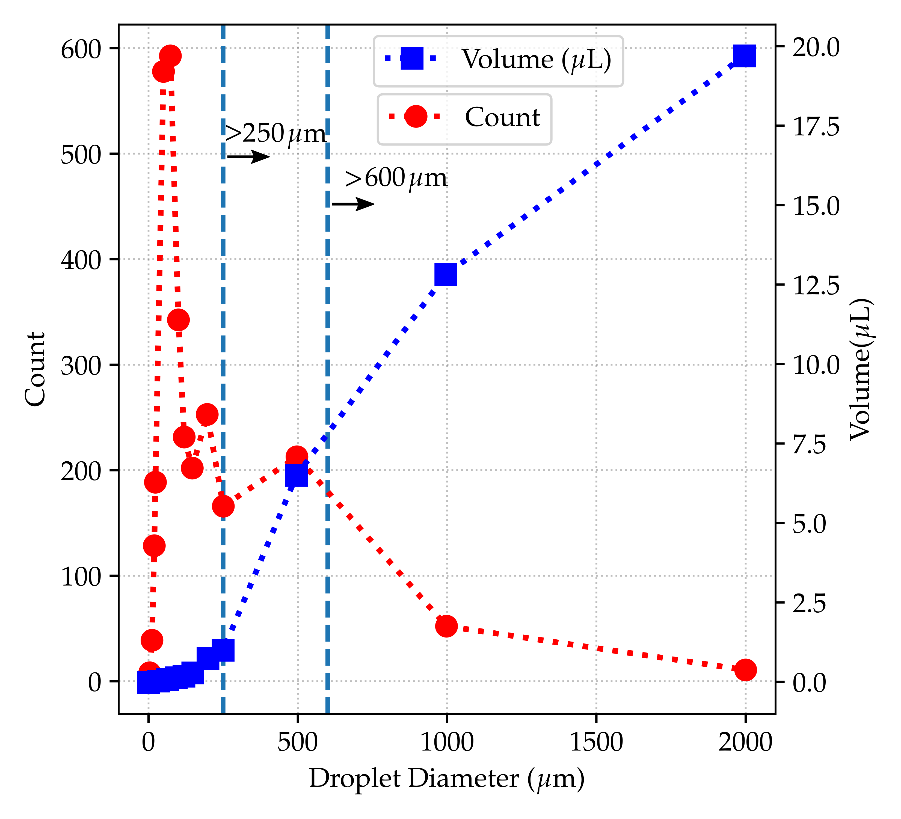

**Fig. S1.** Size and volume distribution of droplets expelled during cough event as given in Duguid (1946) [1]. Two vertical lines with arrows are used to indicate the sizes greater than 250 mm and 600 mm.

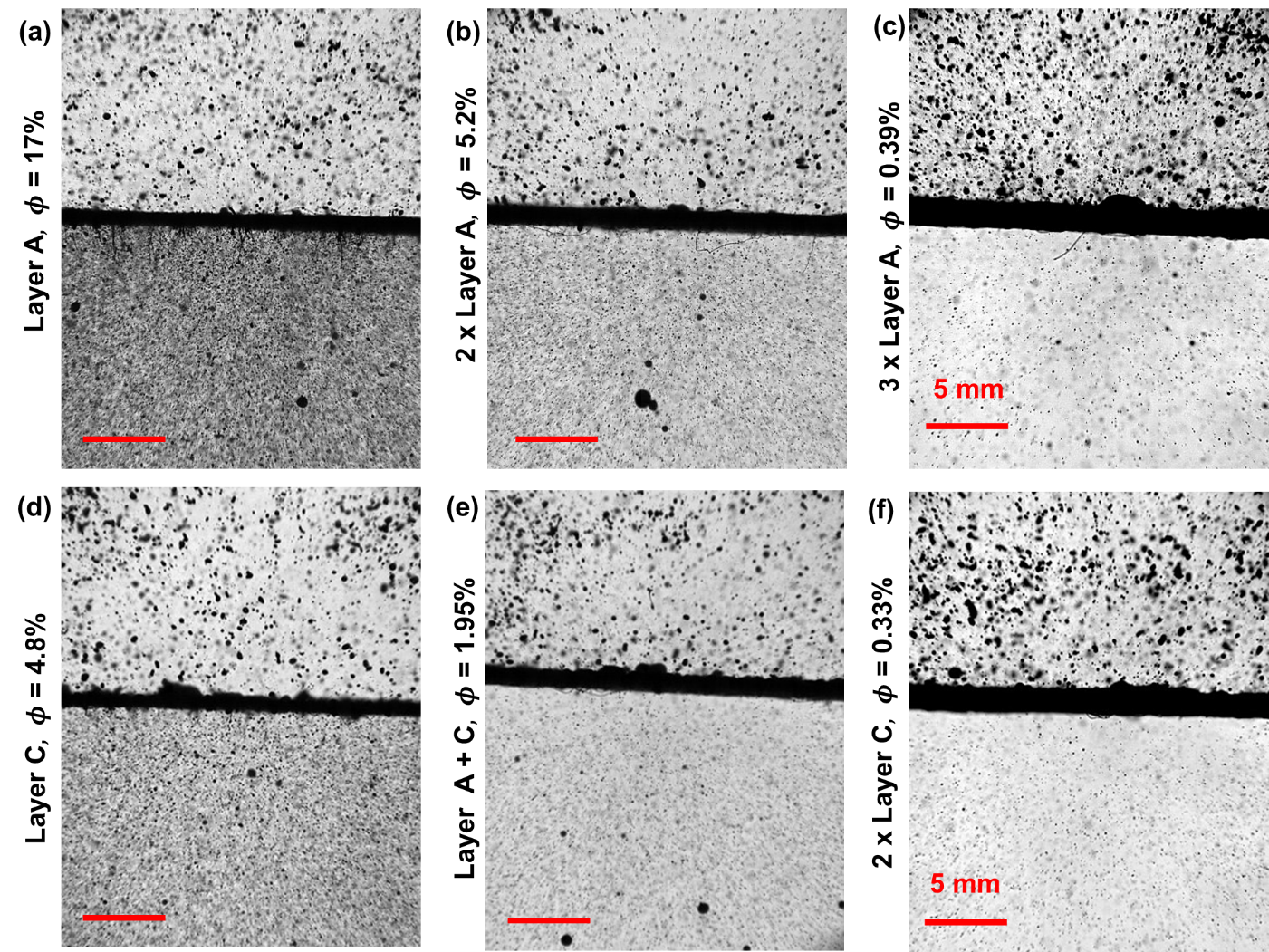

**Fig. S2** The figure shows the spray characteristics and the experimental images of spray impingement on mask surface. **(a-f)** Zoomed in shadowgraphy images of the spray impingement on mask are shown: **(a)** Layer **A**, **(b) 2 x** Layer **A**, **(c)** **3 x** Layer **A**, (**d)** Layer **C**, **(e)** Layer **A + C**, **(f)** **2 x** Layer **C**. The values of porosity ($\boldsymbol{\phi}$) for each sample are shown. All the scale bars represent 5 mm.
